# Dementia Risk Analysis Using Temporal Event Modeling on a Large Real-World Dataset

**DOI:** 10.1101/2023.03.24.23287651

**Authors:** Richard Andrew Taylor, Aidan Gilson, Ling Chi, Adrian D. Haimovich, Anna Crawford, Cynthia Brandt, Phillip Magidson, James Lai, Scott Levin, Adam P. Mecca, Ula Hwang

## Abstract

The objective of the study is to identify healthcare events leading to a diagnosis of dementia from a large real-world dataset. This study uses a data-driven approach to identify temporally ordered pairs and trajectories of healthcare codes in the electronic health record (EHR). This allows for discovery of novel temporal risk factors leading to an outcome of interest that may not otherwise be obvious. We identified several known (Down syndrome RR: 116.1, thiamine deficiency RR: 76.1, and Parkinson’s disease RR: 41.1) and unknown (Brief psychotic disorder RR: 68.6, Toxic effect of metals RR: 40.4, and Schizoaffective disorders RR: 40.0) factors for a specific dementia diagnosis. The associations with the greatest risk for any dementia diagnosis were found to be primarily related to mental health (Brief psychotic disorder RR: 266.5, Dissociative and conversion disorders RR: 169.8), or neurologic conditions or procedures (Dystonia RR: 121.9, Lumbar Puncture RR: 119.0). Trajectory and clustering analysis identified factors related to cerebrovascular disorders, as well as diagnoses which increase the risk of toxic imbalances. The results of this study have the ability to provide valuable insights into potential patient progression towards dementia and improve recognition of patients at risk for developing dementia.

## Introduction

### Background

In the United States, there are currently over 7 million persons living with dementia (PLwD), with nearly 12 million estimated by 2040.[1] PLwD represent a particularly vulnerable population and the increasing number of PLwD are likely to have a disproportionate future impact on the healthcare system.[2, 3] PLwD are known to have higher rates of healthcare utilization including greater emergency department presentations, more hospital admissions, and prolonged hospital courses. Additionally, PLwD have increased risk for morbidity, such as delirium, and falls, as well as higher rates of mortality after healthcare encounters compared to patients without dementia.[4–6] Recognizing and improving care for PLwD has thus become a key national objective with leaders advocating for tailored care practices aimed at enhancing outcomes and care transitions.[7]

### Importance

One key aspect of delivering better dementia care in the future is a firm understanding of what events or sequence of events may be markers for a diagnosis of dementia.[8, 9] While past research has identified key risk factors for cognitive decline, such as age, diabetes, and hypertension[10], few incorporate any notion of time dependency.[11, 12] Trajectory-based risk assessment is a data-driven method used to identify risk factors associated with a specific outcome of interest. Elucidating temporally antecedent risk factors and health event trajectories can significantly enhance our understanding of disease processes or contributory health events, thereby enabling the development of tailored, evidence-based interventions and preventive strategies.[13, 14] Furthermore, by integrating these insights into policy making and healthcare infrastructure, stakeholders can cultivate highly effective and streamlined healthcare systems that are well-prepared to address the mounting challenges associated with the increasing prevalence of dementia. While previous temporal-risk research has used diagnosis trajectories to describe disease progression for non-specific outcomes,[15, 16] there are no studies employing this temporal and trajectory based methodology for dementia or its various subtypes.

### Goals of This Investigation

In the present investigation, we aimed to attain a more comprehensive understanding of the healthcare events in the EHR that precede a dementia diagnosis. Our primary focus was to delineate the temporal associations between these factors and to discern distinct clusters representing diverse dementia trajectories. This research builds upon prior efforts in other areas by employing an unbiased, data-driven methodology to extract potential risk factors from an extensive range of sources, including prior diagnoses, medications, and medical procedures.[17] The trajectories serve as valuable tools for pinpointing critical junctures in the progression of dementia and for identifying patients who may otherwise remain undetected. By enhancing our understanding of the temporal relationships and the diverse trajectories that characterize dementia development, this study contributes to the ongoing efforts to improve early identification, intervention, and overall management of dementia within the healthcare system.

## Methods

### Study Design and Setting

This retrospective observational cohort study focused on patients between 2013 and 2022 seen in the emergency department (ED) at least once across ten sites within a regional healthcare network in the northeastern United States. The sites encompass a geographic area of approximately 650 square miles and closely resemble the overall national population.[8] We included visits for all adult patients *≥*18 years of age. This study followed STROBE reporting guidelines for observational studies.[9] Our institutional review board approved this study and waived the need for informed consent (HIC# 1602017249).

### Data Collection and Processing

Patient demographic and clinical data were extracted from the system-wide electronic health record (Epic, Verona, WI) using a centralized data warehouse (Helix). The initial patient cohort contained information present within the EHR about patients who had at least one ED visit between 2013 and 2022, not just information from that visit alone. The dataset includes procedural instances (Epic Procedural codes), prescription and medications, ICD-10 code diagnoses, and laboratory records. Patients with complete demographic information including age, race, and sex were included. Finally, following previously published methodology, we only included the first instance of an ICD10 code, procedural instance, or medication prescription, and the first instance of an abnormal lab, as it could appear multiple times in a patient’s EHR.[16] With the exception of codes specifying a dementia diagnosis, all ICD10 codes were truncated to three characters.[16] Outpatient medications were mapped to the First DataBank Enhanced Therapeutic Classification System.[18]

### Trajectory Creation

The methodology for trajectory creation follows three steps. First, Fisher exact tests were performed to identify any pairwise combination of ICD10 codes, procedures, medications, or labs that co-occur significantly for a patient (p < 0.05) over the entire study period. Next, a Bernoulli trial was run for each of these identified pairs of the form (C1, C2) to identify if one diagnosis within each pair occurred before the other at a statistically significant rate. Finally, further reduction of these pairs was performed by comparing each patient that temporally followed the codes C1 →C2 sequentially within a cohort of 10,000 other patients who were of the same sex, race, and whose ages fell in the same decade of life.[16] Inclusion of a pair was dependent on the relative risk (RR) of C2 being present and following C1 being greater than one and p ¡0.05 with Bonferroni correction. Specifics of the methodology are outlined by Jensen et al, and in our prior work.[16, 17]

To reduce computational complexity of the future analysis, a breadth first search algorithm was performed on the directed acyclic graph (DAG) constructed from all temporal pairs.[19] Only diagnoses reachable from a dementia diagnosis were included. An additional threshold for inclusion was that 500 patients must traverse the complete trajectory sequentially. To complete the trajectories with a terminal diagnosis of dementia, pairs of codes with overlapping elements were combined (C1→ C2 and C2 →C3 becomes C1 →C2 →C3), such that the terminal diagnosis was a dementia related code. The same methodology of cohort analysis was then performed to control for demographic covariates and produce a RR and p-value for the trajectories.

### Dynamic Time Warping

These identified trajectories were then combined into clusters with overlapping codes using dynamic time warping (DTW).[20] DTW allows for clustering of time-sequenced series with varying lengths based on their components. A technical description of the methodology is provided in the supplements. In summary, each code, procedure, or medication present in a trajectory was embedded in a 256-dimensional space using OpenAI’s GPT-3 model.[21] Distances were then calculated between all pairs of individual trajectories using DTW, and the trajectories were then clustered using these distances. RR for each cluster was calculated in the same way as individual trajectories.

## Results

### Patient Cohort

In total, 442,278 patients seen in a large New England Health System between 2013 and 2022 met inclusion criteria. The average age of the population was 39.0 (std 25.3) years, 60.8% female, 80.9% White and 9.7% Black, and 90.2% Non-Hispanic. Additional sample characteristics are reported in Supplementary Table The dataset includes 34 million procedural instances, 18 million prescriptions, 168 million ICD-10 code diagnoses, and 23 million laboratory records. Patients with incomplete demographic information including age, race, or sex were excluded from the study totalling 24,663.

### Risks from Individual Codes

Analysis began with 12,605 unique diagnoses, labs, procedures, or medications which were present in the past medical history of at least one patient. Within the sample, 79 million pairwise combinations of these codes were present in the past medical history of a patient. After Fisher exact tests for significance, Bernoulli trials for temporality, and cohort matched analysis to control for cofactors, 3.2 million significant directional pairs between any two codes were identified.

Figure 1 shows the 2-dimensional embedding of the codes used in the analysis. Although the overall network is not used directly in the analysis, it provides a qualitative metric to analyze the proximity of codes based on the higher dimensional embeddings generated using GPT-3. These distances play a key role in clustering of the trajectories through DTW.

**Fig. 1.**
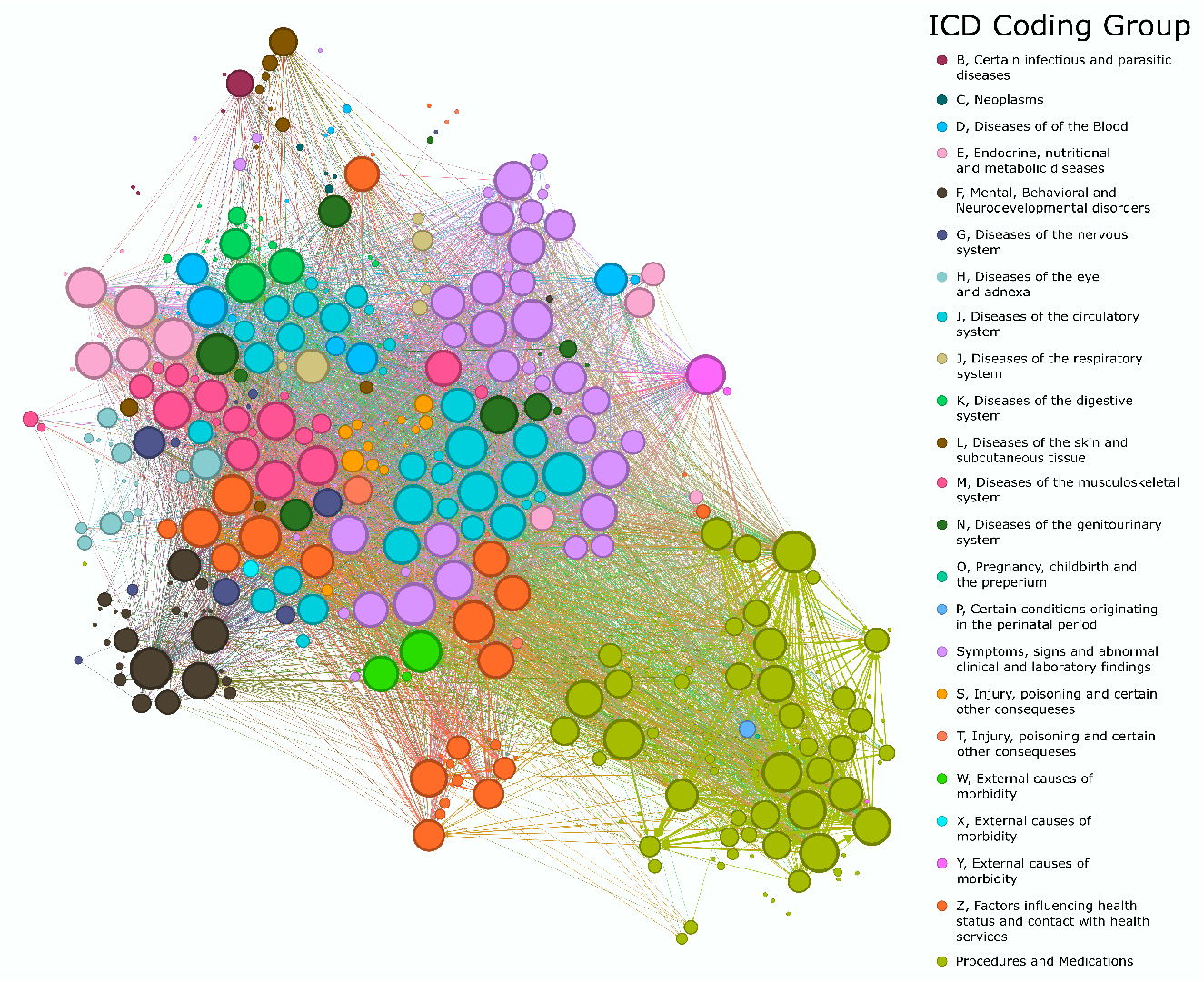
tSNE embedding of the GPT-3 produced feature space of all codes in a trajectory ending in a dementia diagnosis. This figure presents a t-distributed Stochastic Neighbor Embedding (tSNE) visualization of the feature space generated by the GPT-3 model for all codes associated with healthcare trajectories that culminate in a dementia diagnosis. The tSNE embedding technique effectively reduces the high-dimensional data into a two-dimensional representation, enabling a clear visualization of the relationships and patterns among the various codes. In this figure, each point represents a distinct code, and their relative positions in the plot reflect their similarities or dissimilarities in the GPT-3-produced feature space.

Within all identified pairs, 4,472 had a terminal code being a dementia diagnosis. The individual diagnosis with the greatest risk for a dementia diagnosis is Down syndrome which leads to an Alzheimer’s diagnosis with early onset with a RR of 116.1 (95% CI 43.22 - 311.97). The top 25 codes with the greatest predicted increased RR for a specific diagnosis of dementia are shown in Table 1, along with the specific diagnostic code they were found to predict. Supplementary Table 2 shows the top 15 codes which were found to incur the greatest RR for any diagnosis of dementia. Brief Psychotic Disorder was found to correlate with the greatest risk (RR 266.5 95% CI 266.82-268.5). Eight of the top 15 factors were mental health related, with seven diagnoses and one medication use). Five were primarily neurological, consisting of two diagnoses (Parkinson’s and other paralytic symptoms), one symptom (dystonia), one medication category (Drugs primarily affecting the autonomic nervous system), and one procedure (lumbar puncture). Finally two are common during the initial workup of suspected cognitive decline (Brain MRI with 3D Volumetric Analysis, and inpatient consult to Geriatrics).

**Table 1.**
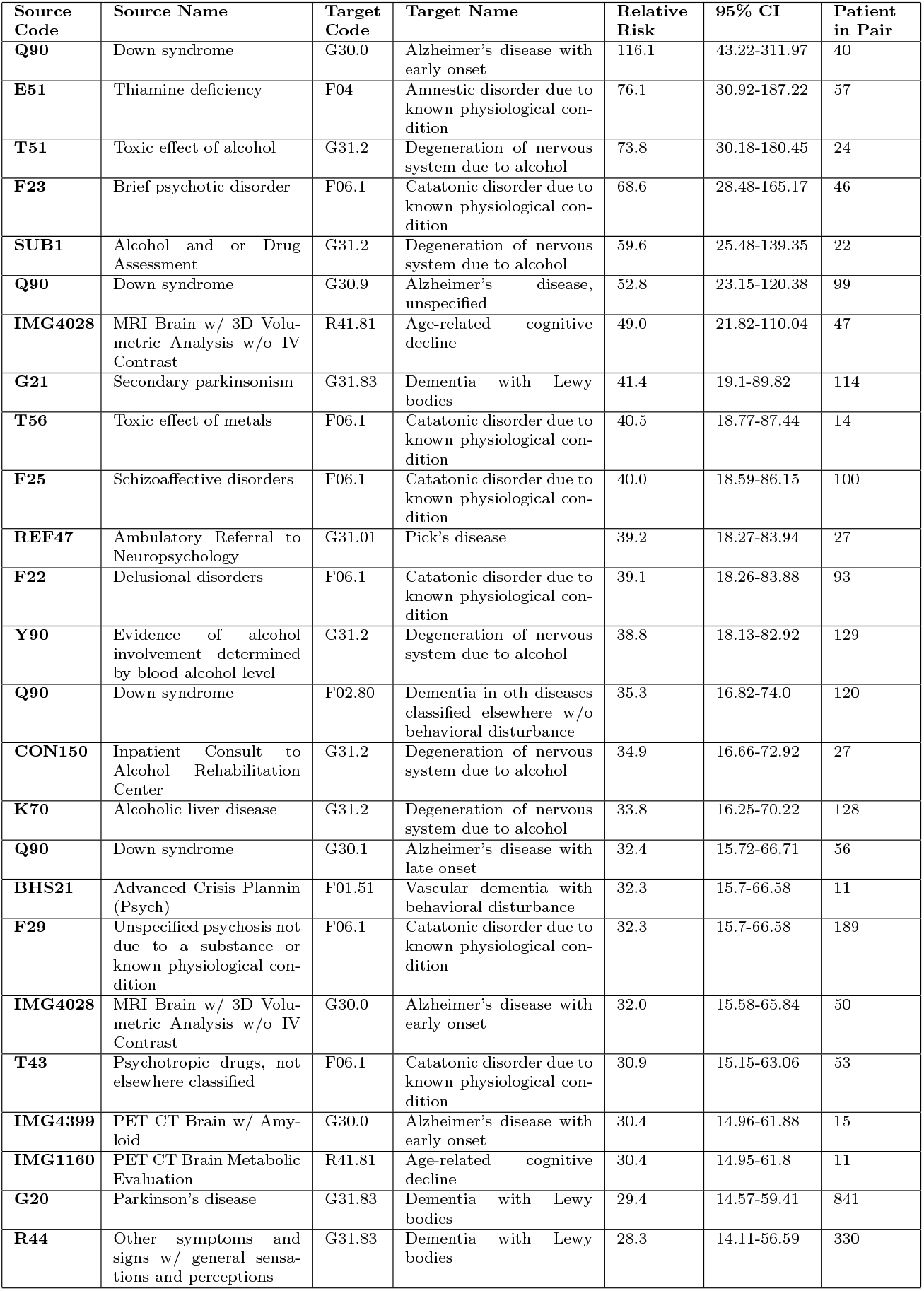
Top 25 Codes by Relative Risk that lead to a specific Dementia Diagnosis

Figure 2 shows the diagnostic codes which lead to any diagnosis of dementia. There is a general negative correlation between patient number and RR, indicating specificity of the association often correlates with a larger RR.

**Fig. 2.**
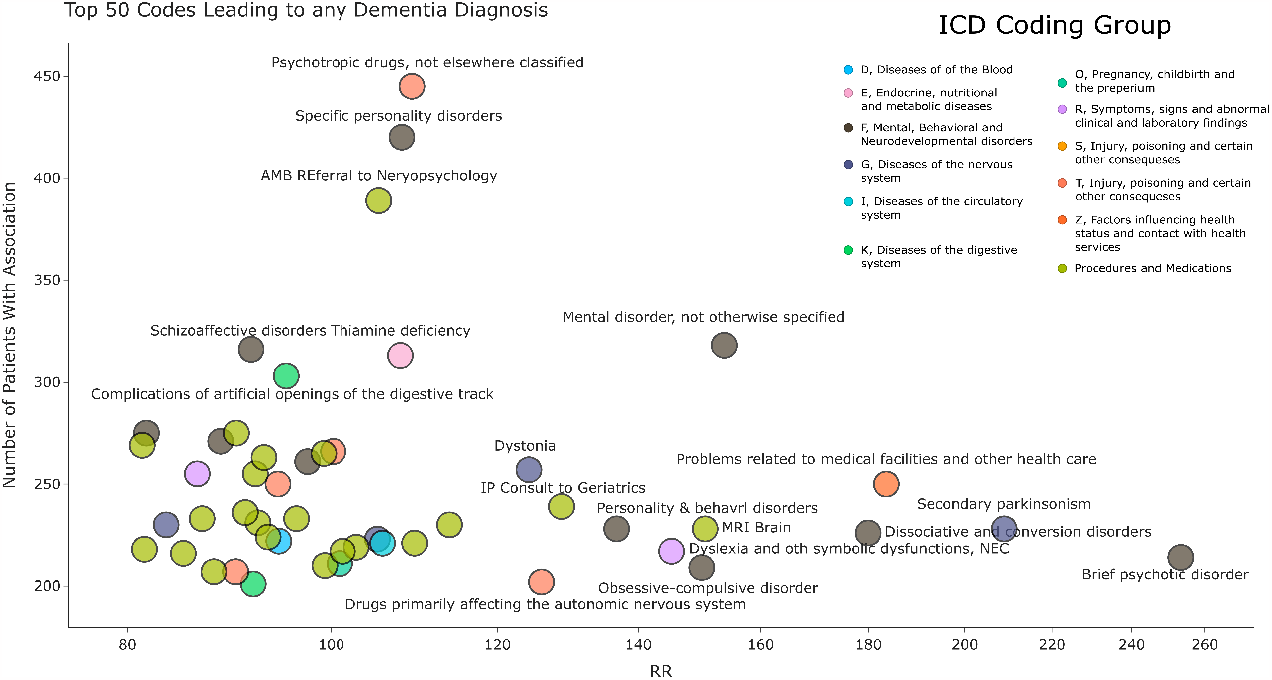
Top 50 codes leading to any dementia diagnosis. The x-axis is the relative risk of the diagnosis for any dementia diagnosis, the y-axis is the number of patients with that association.

### Trajectory Risks

All pairwise combinations of codes outlined in the previous analysis were then combined into trajectories with a cutoff of 500 patients needing to traverse the trajectory in order. RR was calculated through cohort analysis to control for demographic variation in the same way as individual pairs. The top 15 trajectories by RR are shown in Table 2. The top trajectory consisted of a type 2 diabetes diagnosis followed by cerebral infarction, and terminating in Unspecified dementia without a behavioral disturbance (RR 26.4, 95% CI 26.34-26.38). As many of the trajectories with the highest relative risk terminate in F03.90, Unspecified Dementia, we show an additional table (Table 3) with the top trajectories terminating in a vascular dementia diagnosis in order to provide a breadth of results.

**Table 2.** Top 15 Trajectories Terminating in a Dementia Diagnosis

| Codes | Text Description | Relative Risk | 95% CI | Patients in Pair |
| --- | --- | --- | --- | --- |
| E11 → I63 → F03.90 | Type 2 diabetes mellitus → Cerebral infarction → Unspecified dementia without behavioral disturbance | 26.4 | 26.34-26.38 | 520 |
| I10 → F43 → F03.90 | Essential (primary) hypertension → Reaction to severe stress, and adjustment disorders → Unspecified dementia without behavioral disturbance | 25.7 | 25.73-25.77 | 523 |
| E78 → R45 → F03.91 | Disorders of lipoprotein metabolism and other lipidemias → Symptoms and signs involving emotional state → Unspecified dementia with behavioral disturbance | 22.3 | 22.28-22.31 | 596 |
| I10 → L97 → F03.90 | Essential (primary) hypertension → Non-pressure chronic ulcer of lower limb, not elsewhere classified → Unspecified dementia without behavioral disturbance | 22.3 | 22.25-22.27 | 480 |
| E78 → R45 → F02.81 | Disorders of lipoprotein metabolism and other lipidemias → Symptoms and signs involving emotional state → Dementia in other diseases classified elsewhere w/ behavioral disturbance | 22.2 | 22.22-22.25 | 504 |
| J44 → R26 → F03.90 | Other chronic obstructive pulmonary disease → Abnormalities of gait and mobility → Unspecified dementia without behavioral disturbance | 22.1 | 22.1-22.12 | 570 |
| F32 → N18 → F03.90 | Depressive episode → Chronic kidney disease (CKD) → Unspecified dementia without behavioral disturbance | 21.9 | 21.94-21.96 | 585 |
| E78 → I69 → F03.90 | Disorders of lipoprotein metabolism and other lipidemias → Sequelae of cerebrovascular disease → Unspecified dementia without behavioral disturbance | 21.9 | 21.92-21.94 | 616 |
| F32 → Z79 → E87 → F03.90 | Depressive episode → Long term (current) drug therapy → Other disorders of fluid, electrolyte and acid-base balance → Unspecified dementia without behavioral disturbance | 21.7 | 21.7-21.73 | 593 |
| I63 → E87 → F03.90 | Cerebral infarction → Other disorders of fluid, electrolyte and acid-base balance → Unspecified dementia without behavioral disturbance | 21.4 | 21.36-21.39 | 542 |
| J44 → F32 → F03.90 | Other chronic obstructive pulmonary disease → Depressive episode → Unspecified dementia without behavioral disturbance | 21.3 | 21.24-21.26 | 684 |
| I10 → I63 → Z86 → F03.90 | Essential (primary) hypertension → Cerebral infarction → Personal history of certain other diseases → Unspecified dementia without behavioral disturbance | 21.1 | 21.09-21.11 | 591 |
| I10 → I48 → F32 → F03.90 | Essential (primary) hypertension → Atrial fibrillation and flutter → Depressive episode → Unspecified dementia without behavioral disturbance | 21.0 | 21.01-21.04 | 545 |
| I63 → I69 → F03.90 | Cerebral infarction → Sequelae of cerebrovascular disease → Unspecified dementia without behavioral disturbance | 21.0 | 20.99-21.02 | 567 |
| I10 → E78 → F41 → F32 → F03.90 | Essential (primary) hypertension → Disorders of lipoprotein metabolism and other lipidemias → Other anxiety disorders → Depressive episode → Unspecified dementia without behavioral disturbance | 20.9 | 20.85-20.87 | 610 |

**Table 3.** Top 10 Trajectories Terminating in a Vascular Dementia Diagnosis

| **Codes** | **Text Description** | **Relative Risk** | **95% CI** | **Patients in Pair** |
| --- | --- | --- | --- | --- |
| I10 → I67 → F01.50 | Essential (primary) hypertension → Other cerebrovascular diseases → Vascular dementia without behavioral disturbance | 16.2 | 16.2-16.21 | 544 |
| I10 → F05 → F01.50 | Essential (primary) hypertension → Delirium due to known physiological condition → Vascular dementia without behavioral disturbance | 14.4 | 14.37-14.38 | 500 |
| I10 → I63 → F01.50 | Essential (primary) hypertension → Cerebral infarction → Vascular dementia without behavioral disturbance | 11.7 | 11.66-11.67 | 557 |
| E78 → G93 → F01.50 | Disorders of lipoprotein metabolism and other lipidemias → Other disorders of brain → Vascular dementia without behavioral disturbance | 8.7 | 8.71-8.71 | 633 |
| E78 → F41 → F01.50 | Disorders of lipoprotein metabolism and other lipidemias → Other anxiety disorders → Vascular dementia without behavioral disturbance | 5.5 | 5.54-5.54 | 661 |
| E78 → F32 → F01.50 | Disorders of lipoprotein metabolism and other lipidemias → Depressive episode → Vascular dementia without behavioral disturbance | 5.5 | 5.49-5.49 | 835 |
| I10 → M62 → F01.50 | Essential (primary) hypertension → Other disorders of muscle → Vascular dementia without behavioral disturbance | 5.5 | 5.46-5.47 | 528 |
| I10 → F32 → F01.51 | Essential (primary) hypertension → Depressive episode → Vascular dementia with behavioral disturbance | 5.3 | 5.33-5.33 | 567 |
| I10 → E78 → F32 → F01.50 | Essential (primary) hypertension → Disorders of lipoprotein metabolism and other lipidemias → Depressive episode → Vascular dementia without behavioral disturbance | 5.2 | 5.18-5.18 | 679 |
| I10 → F32 → F01.50 | Essential (primary) hypertension → Depressive episode → Vascular dementia without behavioral disturbance | 5.1 | 5.13-5.13 | 953 |

Finally, these trajectories were formed into clusters through DTW. Figure 3 shows three clusters selected as representing unique patient cohorts with significant risk for dementia diagnoses. It is possible for a single patient to be a member of multiple trajectories and clusters. Cluster A represents a cohort with procedures related to nursing/ED evaluation which share trajectories with diagnoses of essential hypertension and hyperlipidemia. The RR for this cohort is 15.76. Cluster B, with a RR of 12.34, again contains essential hypertension and hyperlipidemia, as well as diagnoses of osteoarthritis and unstable gait. Finally Cluster C, is a cohort of patients with ischemic heart disease, chronic heart failure, kidney injury, and hyperlipidemia. The RR of this cluster was 15.76

**Fig. 3.**
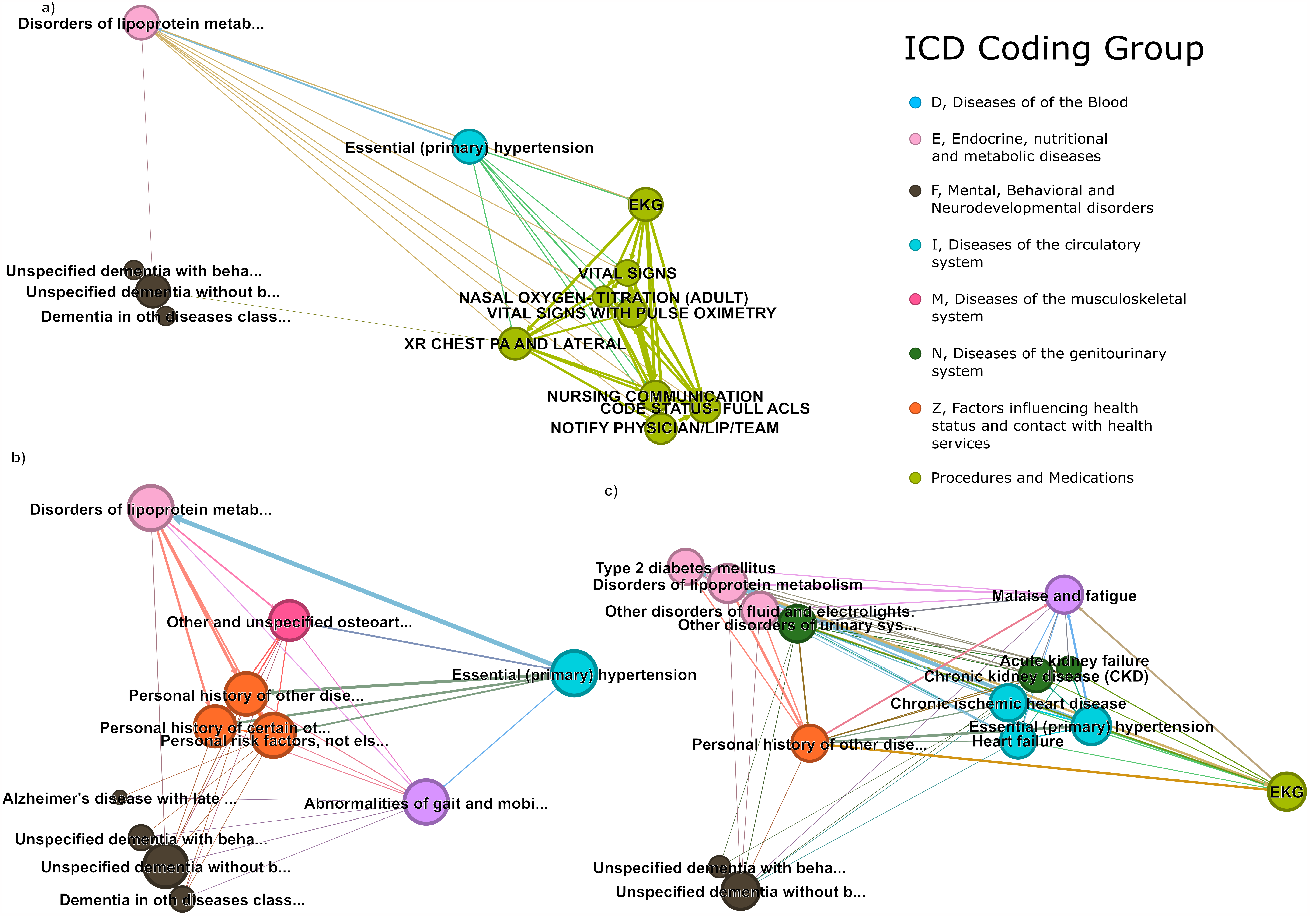
Example Clusters Produced through Dynamic Time Warping (DTW). DTW is a robust technique that allows for the comparison and alignment of time series data, even when the sequences differ in length, scale, or temporal dynamics. It is particularly useful for revealing similarities and differences among the sequences by measuring the optimal alignment between them, taking into account any possible time shifts, expansions, or contractions. Each cluster represents a group of data sequences that exhibit similar patterns or trends, as determined by the DTW analysis.

## Discussion

Improvement of dementia diagnosis and risk stratification is of critical importance as the population continues to age. Unraveling temporally antecedent risk factors and health event trajectories has the potential to improve our understanding of disease processes, paving the way for tailored interventions and preventive strategies. Historically, research concerning the precursors to dementia has primarily focused on the identification of individual factors without considering temporal dependencies. However, recent large-scale, temporally-based methodologies facilitate the unsupervised detection of previously undiscovered risk factors and trajectories, which may not be readily identifiable through traditional approaches. Utilizing these methodologies on a large-scale real-world dataset of patients with dementia, we report several notable findings. Firstly, our results validate established risk factors for dementia or dementia-related illnesses. Secondly, we identify novel temporally antecedent risk factors, including dyslexia and other symbolic dysfunctions (RR:143.4), artificial openings of the digestive tract (RR: 95.1), and specific personality disorders (RR: 145.2). Thirdly, we reveal, for the first time, healthcare event trajectories leading to dementia including progression of alcohol use disorder and degeneration of the nervous system due to alcohol. Lastly, we elucidate intriguing disease trajectory clusters associated with dementia. One cluster demonstrates how intermediate events modify the risk from an initial event, where the risk from common conditions such as hypertension and hyperlipidemia, is modified by nursing examinations and complete emergency department evaluation (Figure 3 Cluster a). Another cluster demonstrates the progression of T2DM to kidney failure and dementia diagnosis (Figure 3 Cluster c).

Numerous individual healthcare events have been associated with dementia. Established risk factors include genetic predispositions (e.g., APOE4 allele and Down syndrome)[22], common medical conditions (e.g., diabetes, hypertension)[23, 24], neurodegenerative disorders, (e.g., Parkinson’s disease, Lewy body dementia)[25], and nutritional deficiencies (e.g., thiamine deficiency)[26]. Our results align with these prior findings and because of the temporality constraint point to deeper causal relationships. Other known associations we confirm include psychotic, schizophrenic and delusional disorders and subsequent catatonia.[27] Additionally, our results align with well-known prior causal mechanisms for dementia subtypes. For example, with vascular dementia, we identify numerous cardiovascular factors (Septic arterial embolism RR: 6.33, Cerebrovascular disorders in diseases classified elsewhere RR: 7.9) which contribute to increased diagnostic risk. Moreover, our analysis identifies the well-known progression to dementia from alcohol use disorder and degeneration of the nervous system due to alcohol. This is also found through both the chronic effects of alcohol, in Toxic effects of Alcohol, as well as acute intoxication, evidence of alcohol involvement determined by blood alcohol level.

In addition to known risk factors, we discover many novel temporal associations. For example, we find dyslexia and other symbolic dysfunction is associated with an increased relative risk (RR:143.4) for all dementia diagnoses. Although there is no known physiologic reason for this association, a dyslexia diagnosis may create barriers that prevent patients from receiving similar levels of cognitive stimulation as their peers. This decreased stimulation may then contribute to a susceptibility for dementia. The dyslexia may lead to the patient having difficulty on screening exams assessing cognitive function, causing an artificially decreased score. A patient having been put on suicide precautions in the past also was associated with increased risk (RR: 96.2). This may be a cofactor of other psychological conditions which were found to be associated with dementia. It may be that this is an early manifestation of a patient’s cognitive decline, not necessarily true suicidal ideation, but the “abnormal” behavior that leads a provider to begin suicide precautions. We also find personality and behavior disorders due to a known physiological condition (RR: 136.7). Although these diagnoses are specifically added with other physiologic conditions in mind as the cause, there is potential that like suicide precautions, this is an indicator of cognitive decline related to dementia. It may be easy to attribute psychiatric changes to known disorders the patient has, rather than considering novel etiologies like dementia. Finally we see two other codes, complications of artificial openings of the digestive system (RR: 95.2), and inpatient consultation with an ostomy nurse (RR: 101.2), related to the same condition. It may be that these conditions point to some psychological underpinning of the causes leading to ostomy bags that increase dementia risk. Or, the need for an increased level of ostomy care, or an increased number of complications may be a first manifestation of cognitive decline where patients have difficulty caring for themselves. A product of the unsupervised temporal nature of the analysis is the identification of precursors to dementia that are actually steps in the evaluation of a patient with suspected cognitive decline. This is most evident with MRI Brain w/ 3D Volumetric Analysis w/o IV Contrast (RR: 153.8) which is likely ordered after the initial identification of cognitive decline in the patient. Trajectories can ameliorate this complication by looking further upstream to the precursors of the brain MRI.

Previous work by Jensen et al. first described the validity of trajectory analysis as a mechanism for novel risk factor identification and the creation of more clinical specific and temporally oriented diagnostic trajectories.[16] Further work by the same group delved into the specific risk factors for sepsis diagnosis.[28] Giannoula et al. then described the utility of dynamic time warping for broad-scale analysis of identified trajectories through clustering.[20] However, this analysis has not been used to identify risk factors for dementia. We have identified trajectories which contain diagnoses related to common chronic multimorbidities, i.e. essential hypertension, hyperlipidemia, diabetes mellitus, cerebrovascular conditions including cerebral infarction and sequelae of cerebrovascular disease, and depressive disorders. Table 4 outlines trajectories with the highest risk for a subsequent vascular dementia diagnosis, most of which contain hypertension or hyperlipidemia as a step in the trajectory.For example one cluster Essential (primary) hypertension→ Disorders of lipoprotein metabolism and other lipidemias →Depressive episode → Vascular dementia (RR: 5.2) contains both hypertension and hyperlipidemia.

The trajectories demonstrate the modulation of the risk factors by the other diagnoses present in the trajectory, and how the use of a single risk factor often oversimplifies the complexity of a patient’s risk. For instance, the first trajectory - hypertension leading to other cerebrovascular diseases and finally dementia - outlines that those patients have progressed to experiencing adverse vascular events (potentially due to the hypertension) rather than having well-controlled hypertension without symptomatology. These patients are then assigned a higher risk than not only than patients who are completely controlled, but also over patients with other comorbidities, such as depression, or muscle disorders.

Clusters represent patients with significant intensities of healthcare and nursing workups. Perhaps this indicates an increased likelihood of diagnosis simply due to an increased quantity of evaluations that the patient receives. Additionally we also show a cluster which contains a prominence of arthritic disorders, something not obviously associated with cognitive function, but nonetheless provides prognostic value. Finally, we show a cluster which contains cardiorenal comorbidities. Both the trajectories and clusters with the highest risk often contain some of the most common diagnoses present in an older population such as hypertension, hyperlipidemia, depression, and chronic obstructive pulmonary disease. Further research is needed to understand the potential causative nature of these conditions on an eventual dementia diagnosis. However, the constraints of the temporality of the trajectories in our analysis provides initial insight into the timing of the terminal diagnosis. As the maximum time between sequential codes was constrained to five years within the analysis, the trajectories outline the rapid addition of comorbidities for patients. Additionally, the trajectories often contain both chronic cardiometabolic/endocrine disorders such as previously mentioned hypertension and hyperlipidemia, as well as mental health diagnoses, including depression and anxiety. This may point to a synergistic effect of prior diagnoses leading to an eventual dementia diagnosis. The innovative prognostic data delineated in this study has the potential to impact both clinical and research areas. The integration of an individual’s past medical history into trajectory-based analyses, as opposed to relying solely on single associations, enables a more precise determination of risk during clinical encounters. This comprehensive approach has the potential to enhance our understanding of the complex interplay of factors contributing to dementia risk. Incorporating this trajectory-based analysis into EHRs could facilitate automated alerts for healthcare providers, signaling the need for further assessment or intervention in patients with elevated dementia risk. Consequently, this advanced prognostic information may contribute to reducing the under-recognition of dementia and mitigating adverse outcomes associated with delayed or missed diagnoses. In the research domain, the application of these novel prognostic indicators can inform the design and execution of targeted studies. By stratifying populations based on risk trajectories, researchers can focus on specific subgroups of patients, enabling a more granular exploration of the underlying mechanisms driving dementia progression. Moreover, this approach can guide the development of tailored interventions and inform the selection of appropriate endpoints in clinical trials, ultimately leading to more effective therapeutic strategies and improved patient outcomes.

## Limitations

This study has several limitations. Notably, this analytical approach does not establish a causal relationship but rather identifies correlations between prior healthcare information and subsequent dementia diagnosis. Despite this limitation, the clinical application of the findings remains relevant. The correlations are derived from a sizable cohort, exhibiting high statistical significance upon final analysis, thereby substantiating the utility of these associations, trajectories, or clusters as predictive tools, irrespective of established causation. Furthermore, several well-known causal relationships, such as Down syndrome and Alzheimer’s disease or thiamine deficiency and amnestic disorders, lend credibility to the identified associations. It is essential to consider that this study is limited to patients who had at least one ED visit and is based on data from a single institution, which may affect the generalizability of the findings. The temporal ordering of specific trajectories could be influenced by diagnostic preferences and clinical pathways within the healthcare system utilized for the analysis. The choice between two similar yet distinct codes, such as G30.0 (Alzheimer’s disease with early onset) and G30.9 (Alzheimer’s disease, unspecified), might be shaped by factors like practice setting and clinician training, potentially leading to discrepancies in the identified associations.

## Conclusions

In conclusion, we have not only described both novel individual and sequence based temporal associations for dementia, but we have quantified their significance using advanced trajectory analysis. This work provides providers additional resources to screen patients for their risk of cognitive disorders. Additionally, it provides the starting points for further research into each identified risk factor, and suggests targets to reduce dementia risk. Further efforts are needed to evaluate the real-world reduction of missed dementia screening that this research can provide.

## Data Availability

All data produced in the present study are available upon reasonable request to the authors

