## Supplements for "Dementia Risk Analysis Using Temporal Event Modeling on a Large Real-World Dataset"

**Supplement:**

**Supplementary Table 1:**

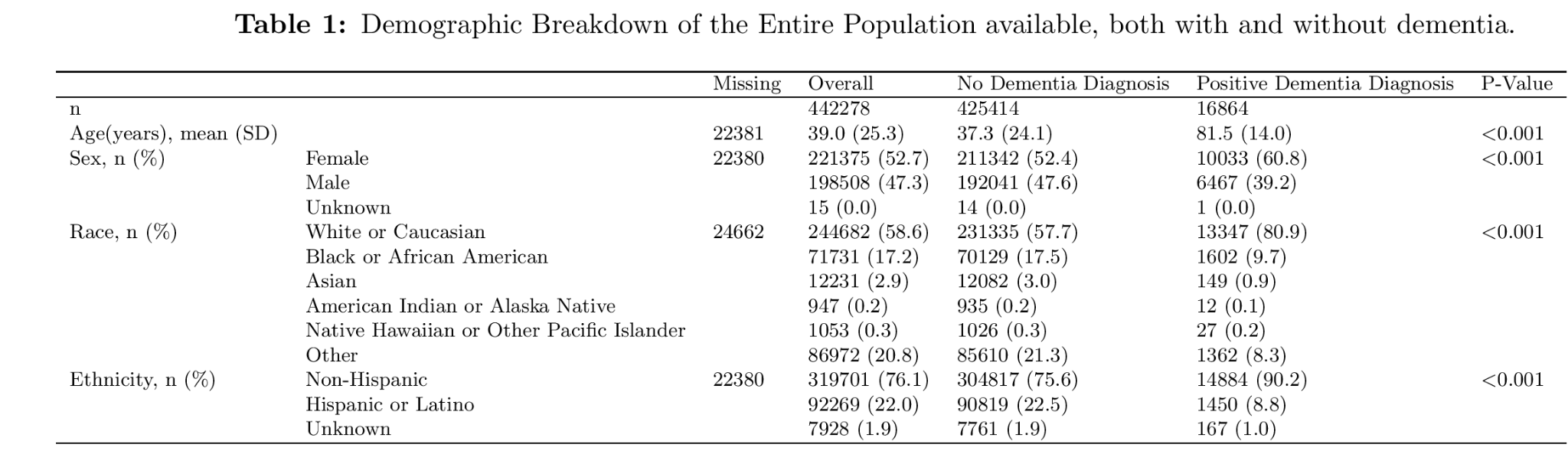

**Supplementary Table 2:** Top 15 Codes by Relative Risk that lead to any dementia diagnosis.

| **Source Code** | **Source Name** | **Relative Risk** | **95% CI** | **Patient in Pair** |
| --- | --- | --- | --- | --- |
| F23 | Brief psychotic disorder | 266.5 | 266.82-268.5 | 214 |
| G21 | Secondary parkinsonism | 200.2 | 199.93-200.96 | 228 |
| Z75 | Problems related to medical facilities and other health care | 181.0 | 180.81-181.69 | 250 |
| F44 | Dissociative and conversion disorders | 169.8 | 169.92-170.75 | 226 |
| F99 | Mental disorder, not otherwise specified | 158.8 | 158.58-159.22 | 318 |
| IMG4028 | MRI Brain w/ 3D Volumetric Analysis w/o IV Contrast | 149.5 | 149.43-150.07 | 228 |
| F07 | Personality & behavioral disorders due to known physiological condition | 145.2 | 145.1-145.72 | 228 |
| R48 | Dyslexia and other symbolic dysfunctions, not elsewhere classified | 143.4 | 143.25-143.87 | 217 |
| F42 | Obsessive-compulsive disorder | 142.2 | 142.13-142.84 | 209 |
| CON55 | IP Consult to Gerontology | 125.2 | 125.09-125.59 | 239 |
| T44 | Drugs primarily affecting the autonomic nervous system | 124.8 | 124.85-125.37 | 202 |
| G24 | Dystonia | 121.9 | 121.75-122.21 | 257 |
| PRO88 | Lumbar Puncture | 119.0 | 119.0-119.48 | 230 |
| G83 | Other paralytic syndromes | 109.4 | 109.38-109.81 | 223 |
| T43 | Psychotropic drugs, not elsewhere classified | 108.9 | 108.92-109.19 | 445 |

**Dynamic Time Warping**

DTW is completed in two steps, calculating the distances between all pairwise combinations of trajectories, and second, clustering those trajectories using the distance metric from the initial step.^16^

The distance between two trajectories of the form${traj}_{one}= \{C_{1a}\to C_{2a}\to...\to C_{Na}\}$ and ${traj}_{two}= \{C_{1b}\to C_{2b}\to...\to C_{Mb}\}$ is given by the matrix D(N,M) where each entry in D is defined as:

$$D(N, M) = Distance(n,m) + min(D(n-1,m), D(n, m-1), D(n-1,m-1))$$

$Distance(n,m)$ is the distance between the n^th^ code of traj_one_ and the m^th^ code of traj_two_. Prior implementations of DTW have utilized the hierarchical structure of the ICD9 coding system to construct a well defined distance calculation between codes. However, given the heterogeneity of lexicons used in our implementation, an alternative distance metric was needed. To calculate distances between codes, we embedded each code using OpenAI’s GPT-3 embedding model. The model used was *text-embedding-ada-002*.^23^ The plaintext name for each code, mapped from the lexicon that it originated (ICD10, CPT, RxNorm, etc) was then queried using the OpenAI API. 256-dimensional embeddings were generated for each code queried. These embeddings were then used in the dynamic time warping algorithm.

Clusters were formed in an unsupervised manner after selection of a maximum separation distance. An initial trajectory is assigned to an arbitrary cluster. Each following trajectory is then compared to the average embedding of each cluster created up until that point. If the minimum distance to any cluster is below the maximum similarity distance, then the trajectory is assigned to that cluster, otherwise a new cluster is formed containing only that trajectory. The maximum separation distance was selected manually to balance adequate merging of trajectories in order to form clusters with interpretable themes without the clusters becoming overly heterogeneous. The RR for each cluster was calculated using a similar formula to that of the trajectory RR .For example, consider a population of 10,000 patients and a cluster defined with two trajectories $\{C_{1}\to C_{2}\to C_{3}\to C_{4}\}$ and $\{C_{5}\to C_{3}\to C_{4}\}$, where 100 patients in the population experience C_4_. If 250 patients in the population traverse one of or both of the trajectories up until C_4_ and of those 250 patients, 50 subsequently experience C_4_ the RR of C_4_ for the cluster is:

$$RR = \frac{(50/250)}{(100/10,000)}=20$$
